# MAIT cell activation and dynamics associated with COVID-19 disease severity and outcome

**DOI:** 10.1101/2020.08.27.20182550

**Authors:** Tiphaine Parrot, Jean-Baptiste Gorin, Andrea Ponzetta, Kimia T. Maleki, Tobias Kammann, Johanna Emgârd, André Perez-Potti, Takuya Sekine, Olga Rivera-Ballesteros, the Karolinska COVID-19 Study Group, Elin Folkesson, Olav Rooyackers, Lars I. Eriksson, Anna Norrby-Teglund, Hans-Gustaf Ljunggren, Niklas K. Björkström, Soo Aleman, Marcus Buggert, Jonas Klingström, Kristoffer Strålin, Johan K. Sandberg

## Abstract

Severe COVID-19 is characterized by excessive inflammation of the lower airways. The balance of protective versus pathological immune responses in COVID-19 is incompletely understood. Mucosa-associated invariant T (MAIT) cells are antimicrobial T cells that recognize bacterial metabolites, and can also function as innate-like sensors and mediators of antiviral responses. Here, we investigated the MAIT cell compartment in COVID-19 patients with moderate and severe disease, as well as in convalescence. We show profound and preferential decline in MAIT cells in circulation of patients with active disease paired with strong activation, as well as significant MAIT cell enrichment and pro-inflammatory IL-17A bias in the airways. Unsupervised analysis identified MAIT cell CD69^high^ and CXCR3^low^ immunotypes associated with poor clinical outcome. MAIT cell levels normalized in the convalescent phase, consistent with dynamic recruitment to the tissues and subsequent release with disease resolution. These findings indicate that MAIT cells are engaged in the immune response against SARS-CoV-2 and suggest their involvement in COVID-19 immunopathogenesis.

**One sentence summary:** MAIT cells are strongly activated by SARS-CoV-2 infection in a manner associated with disease severity and outcome, they decline in blood, are enriched in the airways as a prominent IL-17A expressing subset, and dynamically recover in circulation during convalescence.

## Introduction

Severe acute respiratory syndrome (SARS) Coronavirus-19 (SARS-CoV-2) causes viral pneumonia and coronavirus disease 2019 (COVID-19), which in some individuals progresses to acute respiratory distress syndrome (ARDS) characterized by aggressive inflammatory responses in the lower airways (reviewed in (*1*)). Severe COVID-19 is not only due to direct effects of the virus, but also in part to a misdirected host response with complex immune dysregulation of both innate and adaptive immune and inflammatory components (*2, 3*). The COVID-19 pandemic has been met with an unprecedented research effort by academia as well as the pharmaceutical industry. Nevertheless, by mid-2020 many aspects of COVID-19 immunopathogenesis still remain poorly characterized.

The majority of T cells respond in an adaptive fashion to peptide antigens governed by MHC-restriction, and the role of CD8 and CD4 T cell responses against COVID-19 has recently been demonstrated (*4-10*). However, the T cell compartment also encompasses several unconventional invariant T cell subsets that have innate-like functions (*11*). Mucosa-associated invariant T (MAIT) cells represent 1-10% of T cells in the circulation, have strong tissue homing characteristics and are particularly abundant in the liver and lung (reviewed in (*12*)). MAIT cells are activated by TCR recognition of microbial vitamin B_2_ (riboflavin) metabolites from a range of microbes presented by MHC-Ib-related protein 1 (MR1) molecules (*13*). However, some MAIT cell functions can be activated or co-activated by cytokines such as IL-18 and type I IFNs (*14, 15*). MAIT cells rapidly produce IFNγ, TNF, and IL-17, and mediate effective cytolytic function dependent on granzyme B (GrzB) (*16-18*). This broad effector profile contributes to the role of MAIT cells in the protection against bacterial pulmonary infections (*19-21*), where MAIT cells have a role in recruiting adaptive T cells to the lung (*22*). MAIT cells in mucosa have an IL-17/22-biased functional profile distinct from that of circulating cells (*23*), and can also be pro-fibrogenic in chronic inflammatory disease (*24*).

Emerging evidence indicates that MAIT cells are innate-like sensors of viral infection; Human MAIT cells are activated in response to several RNA viruses (*25, 26*), expand during acute stages of HIV-1 infection (*27*), and may have a protective role in influenza virus infection as deduced from studies in murine models (*28*). Based on these distinct innate-like, tissue homing and pro-inflammatory characteristics of MAIT cells, we set out to study these cells in COVID-19. Our findings indicate that MAIT cells are engaged in the immune response against SARS-CoV-2 and suggest their possible involvement in COVID-19 immunopathogenesis.

## Results

### Profound MAIT cell decline in the circulation and enrichment in the airways of COVID-19 patients

With the aim to study COVID-19 immunopathogenesis, 24 patients were prospectively recruited after admission to the Karolinska University Hospital as part of the Karolinska COVID-19 Immune Atlas project (*10, 29*). Inclusion and exclusion criteria defined two groups of acute patients with moderate (AM) or severe (AS) disease matched with respect to other patient characteristics (Table S1). Fourteen matched healthy donors (HD) who were SARS-CoV-2 IgG seronegative and symptom-free at time of sampling were included as controls. To minimize inter-experimental variability and batch effects, all samples were acquired, processed, and analyzed fresh during three consecutive weeks in the spring of 2020 at the peak of the COVID-19 pandemic in Stockholm, Sweden.

To study changes in conventional and unconventional T cell subsets in patients with AM and AS COVID-19 disease, a 22-parameter flow cytometry panel was designed to evaluate frequency, activation, homing and functional phenotypes of MAIT cells, invariant natural killer T (iNKT) cells, CD4 and CD8 double negative T (DNT) cells (*30*), γδ T cells as well as conventional CD4 and CD8 T cells (Fig. S1A). We initially analyzed the whole data set through an unsupervised approach using Uniform Manifold Approximation and Projection (UMAP) analysis on CD3^+^ single live events in all patients and controls (n=38). Projection of defining markers allowed visualization of the location of distinct T cell subsets on the UMAP topography (Fig. 1A), which was confirmed using manual gating. Projecting data from HD, AM and AS subjects separately, revealed a clear difference between patients and controls with severe reduction in the distinct topography defined by the MR1-5-OP-RU tetramer suggesting loss of MAIT cells in COVID-19 (Fig. 1B). The profound decline in MAIT cell percentage (Fig. 1C), and absolute counts (Fig. 1D), in COVID-19 patients was confirmed by manual gating. The absolute count decline extended to conventional CD4 and CD8 T cell subsets, and DNT cells, while iNKT cells and γδ T cells were largely unchanged. However, the MAIT cell lymphopenia was distinct in its severity, and was pronounced already in the AM group where loss of overall T cell subsets was not significant. The circulating MAIT cell pool comprises three subsets expressing CD8, CD4, or DN displaying some functional differences (*31*). The numerical decline of the DN MAIT cell subset was more marked than that of the main CD8^+^ MAIT cell population (Fig. S1B).

**Fig. 1.**
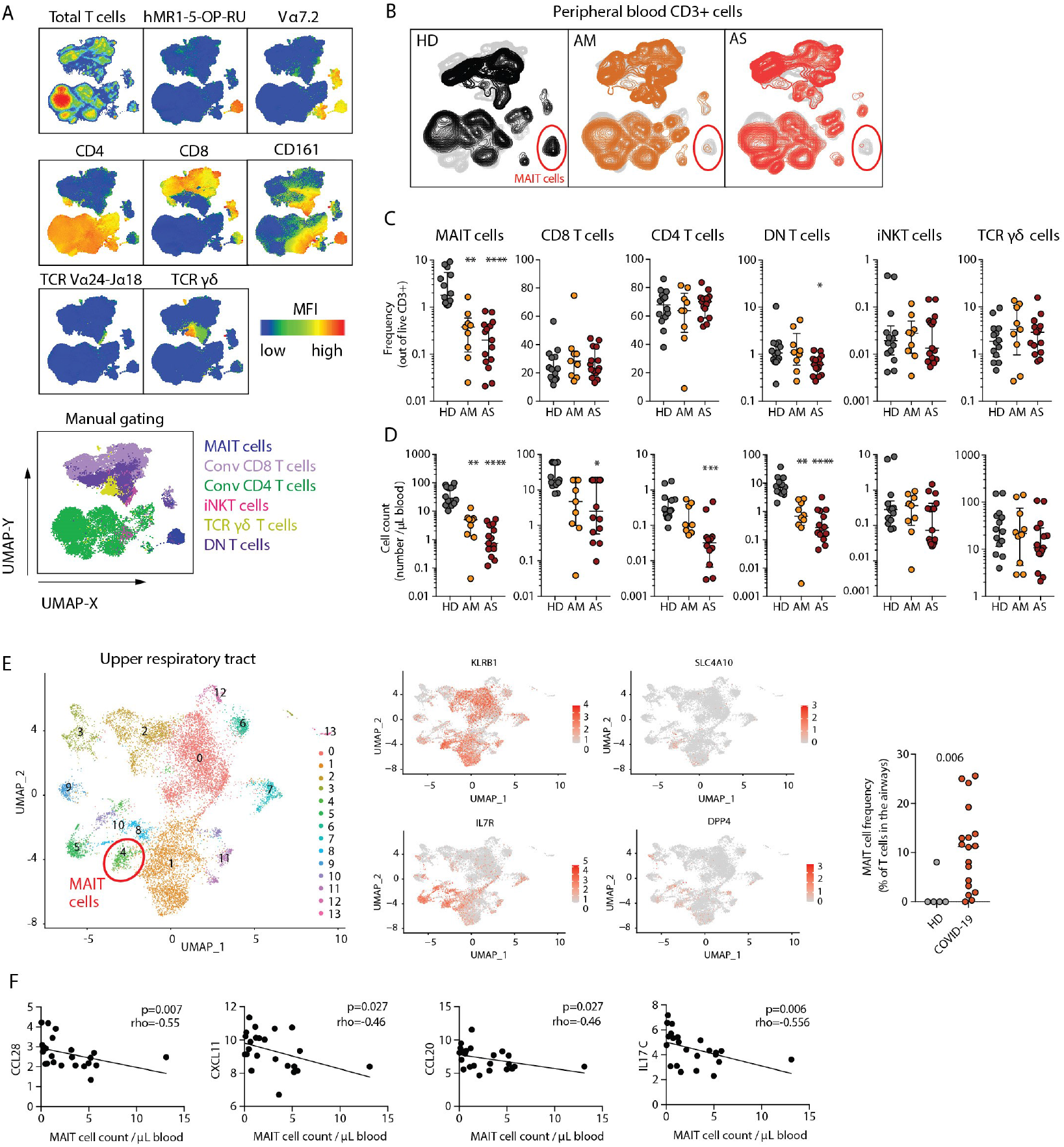
Profound and preferential decline of MAIT cells in blood of patients with COVID-19. **(A)** UMAP plots of total live CD3 T cells in peripheral blood showing the expression of the indicated markers. Bottom: UMAP plots of total live CD3 T cells overlaid with the immune subsets identified by manual gating. **(B)** UMAP plots of total live CD3 T cells in peripheral blood colored according to the patient group: healthy donors (HD, n=14), acute moderate (AM, n=9) and acute severe (AS, n=15). 20,000 CD3 T cells per patients were down-sampled, barcoded according to the patient group and concatenated. The red circle highlights the MAIT cell compartment. **(C)** Dot plots illustrating the relative frequency (median ± IQR), and **(D)** the absolute counts (median ± IQR) of the indicated T cell subsets in peripheral blood. Each dot represents one donor. Non-parametric Kruskal-Wallis test and Dunn’s post-hoc test were used to test for statistical differences between patient groups. *p<0.05, **p<0.01, ***p<0.001, ****p<0.0001. **(E)** Left: UMAP plot displaying the 13 clusters identified based on gene expression levels of non-B lymphocytes in nasopharyngeal swabs (n=4 healthy donors, and n=19 COVID-19 patients). Middle: UMAP plots of the canonical transcripts KLRB1, SLC4A10, IL7R and DPP4 used for MAIT cell identification. Right: graph showing the relative abundance of the MAIT cell containing cluster (cluster 4) in nasopharyngeal swabs from HD and COVID-19 patients. Horizontal bars indicate medians. Each dot represents one donor. Non-parametric Mann-Whitney U test was used to test for significant differences between groups. **(F)** Spearman correlations between the absolute count of MAIT cells and cytokine and chemokine serum levels in COVID-19 patients (n=24). The spearman correlation coefficient (rho) and the associated calculated p-value (p) are indicated on each graph.

Analysis of a publicly available single cell RNA sequencing (scRNAseq) data set on nasopharyngeal samples (*32*), allowed identification of MAIT cells in the upper respiratory tract of COVID-19 patients and healthy control subjects using a combination of MAIT cell defining transcripts (KLRB1, SLC4A10, IL7R and DPP4) (Fig. 1E). Interestingly, the scRNAseq data indicated that MAIT cells were highly enriched within T cells infiltrating the airways of COVID-19 patients as compared to controls (Fig. 1E), consistent with the profound decline of the circulating MAIT cell pool in COVID-19 disease and a possible recruitment to this site. This pattern was corroborated by analysis of MAIT cells in a second published scRNAseq data set on bronchoalveolar lavage (BAL) fluid (*33*) (Fig. S1C). The interpretation that MAIT cells are recruited to the airways in COVID-19 was further supported by inverse correlations between MAIT cell counts and serum levels of CCL20 and CXCL11 in our cohort (Fig. 1F), the receptors for which (CCR6 and CXCR3, respectively) MAIT cells express at high levels (*34*). Furthermore, MAIT cell counts were inversely correlated with IL-17C levels in plasma, supporting a possible link between MAIT cell recruitment and lung epithelium inflammation (Fig. 1F). These soluble factors were all increased in serum of COVID-19 patients compared to HD (Fig. S1D). Together, these findings identify a pattern of profound and preferential decline of circulating MAIT cells in COVID-19, and suggest that this is at least partly caused by recruitment of MAIT cells to the airways.

### Display items: 4 figures, 5 supplementary figures, and 3 supplementary tables

Next, we were interested to analyze MAIT cell characteristics in the context of COVID-19. Unsupervised analysis of MAIT cell flow cytometry phenotypes in all Atlas cohort patients and controls (n=38) revealed a pattern of enhanced CD69 expression and diminished CXCR3 expression in both AM and AS COVID-19 patients (Fig. 2A, 2B). The activated CD69^high^ phenotype was shared between AM and AS COVID-19 patients, although AS patients had slightly higher levels of GrzB and Ki67 than AM patients (Fig. 2C). Expression levels of PD-1, IL-7R, CXCR6, Granzyme A (GrzA), and CD56 were similar between HD, AM and AS groups (Fig. S2A). Activation patterns were largely shared between CD8^+^ and DN MAIT cell subsets, although the CD8^+^ MAIT cell pool showed somewhat higher levels of activation (Fig. S2B). The patterns with CD69 upregulation and CXCR3 downregulation were more pronounced in MAIT cells as compared to conventional CD4 and CD8 T cells (Fig. S2C). Correlation analyses of the MAIT cell phenotype data set (Fig. 2D) indicated that activation levels reflected by CD69 expression were inversely linked with CXCR3 expression (Fig. 2E), and MAIT cell percentages were directly correlated with CXCR6 expression (Fig. 2F).

**Fig. 2.**
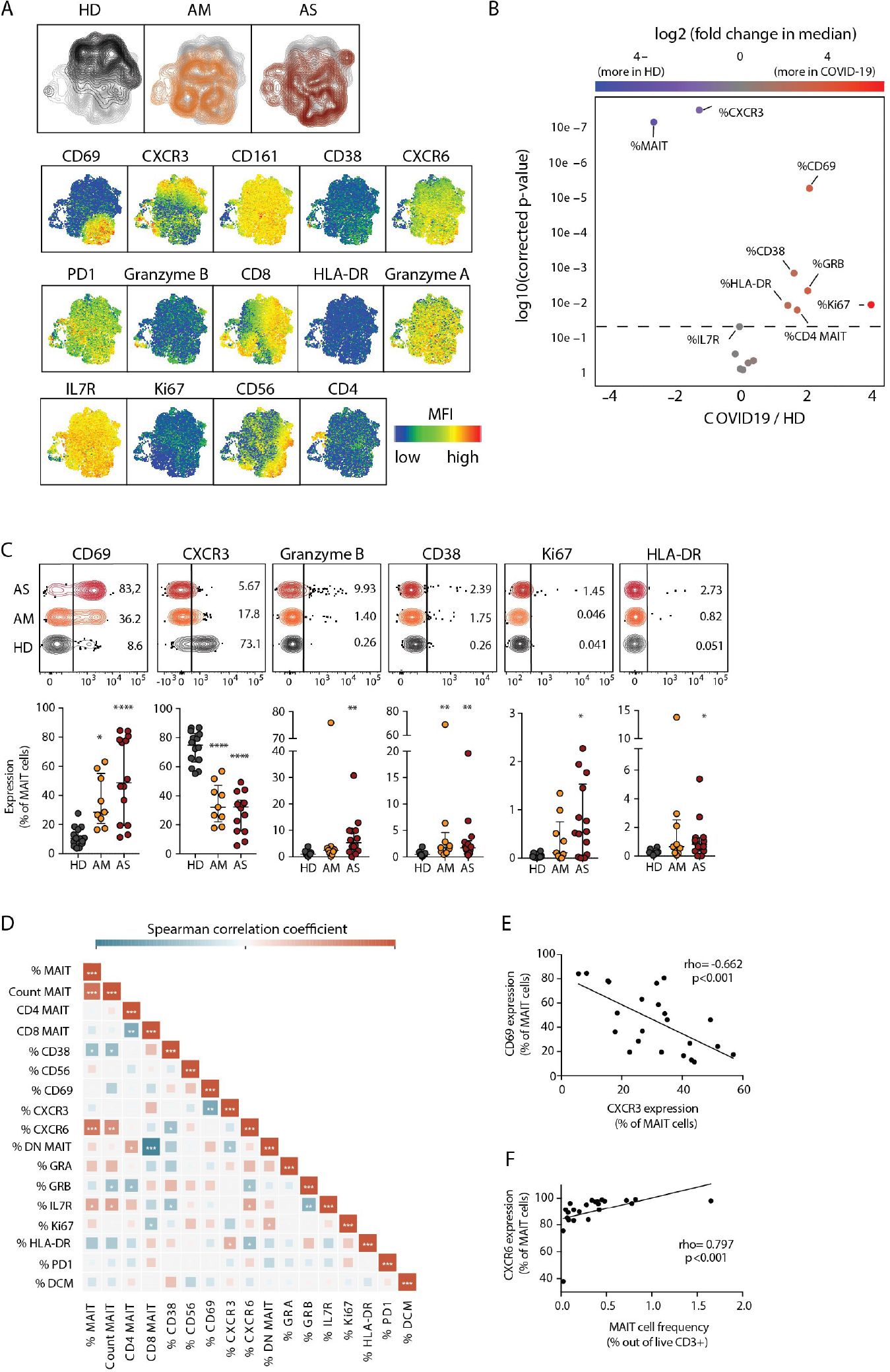
MAIT cell activation and decreased CXCR3 expression in COVID-19. **(A)** Top: UMAP plots of MAIT cells showing the clustering of MAIT cells by patient group. To generate the UMAP, a maximum of 200 MAIT cells per patients were down-sampled, labeled according to patient group and concatenated. Bottom: UMAP plots of MAIT cells showing the expression of the indicated marker. **(B)** Volcano plot showing the log_2_ (fold change) in median expression of the indicated markers on MAIT cells between healthy donors (HD, n=14) and COVID-19 patients (n=23). Significantly up-regulated or downregulated markers (p<0.05) are shown in red and blue, respectively. P-values were calculated using Wilcoxon ranked exact test and adjusted to a FDR of 5% using the Benjamini–Hochberg method. **(C)** Top: Illustrative concatenated flow cytometry plots showing the percentage of expression of the indicated phenotypic markers on MAIT cells by patient group. Bottom: dot plots (median ± IQR) showing the percentage of expression of the indicated markers on MAIT cells in HD (n=14), acute moderate (AM, n=9) and acute severe (AS, n=14) COVID-19 patients. Non-parametric Kruskal-Wallis test and Dunn’s post-hoc test were used to detect significant differences between groups. *p<0.05, **p<0.01, ***p<0.001, ****p<0.0001. **(D)** Heatmap displaying pairwise Spearman correlations between MAIT cell phenotypic parameters in COVID-19 patients. Color indicates the strength of the correlation. *p<0.05, **p<0.01, ***p<0.001. **(E)** Spearman correlation between CXCR3 and CD69 expression on MAIT cells in COVID-19 patients. **(F)** Spearman correlation between MAIT cell frequency and CXCR6 expression on MAIT cells in COVID-19 patients. **(E, F)** The spearman correlation coefficient (rho) and the associated calculated p-value (p) are indicated on the graph.

Analysis of the scRNAseq data set on nasopharyngeal samples (*32*), allowed characterization of the transcriptional profile of MAIT cells in the airways of COVID-19 patients (Fig. S3). The CD69^+^CXCR3^-^phenotype of MAIT cells in peripheral blood was reproduced at the transcriptional level in the airways. Strikingly, the transcriptional profile indicated that MAIT cells were the main subset of airway T cells expressing *IL17A*. This profile was paired with expression of *TNF*, and an apparent lack of *IFNG* and *GZMB* transcripts. Together, these results indicate that the residual circulating MAIT cell pool is highly activated with lowered CXCR3 expression; a finding reproduced in the airways together with an *IL17A^+^* transcriptional profile.

### MAIT cell characteristics associated with plasma viremia and disease outcome

We next investigated if any characteristics of the MAIT cell compartment could be associated with the subsequent clinical outcome. Of the 24 patients sampled for the Karolinska COVID-19 Immune Atlas cohort and subjected to MAIT cell analyses, 20 recovered and were discharged while four died in the hospital. Projection of the deceased patients on unsupervised UMAP analysis of MAIT cells from all patients and controls (n=38) revealed a distinct distribution pattern (Fig. 3A, left plot). PhenoGraph clustering revealed four MAIT cell clusters overrepresented in the four patients who later died of COVID-19 (Fig. 3A). These PhenoGraph clusters were characterized by extraordinarily high CD69 expression and low or very low levels of CXCR3 (Fig. 3B and Fig. S4). Interestingly, within the entire Atlas patient group CD69 was higher in patients with detectable plasma viremia (Fig. 3C and Fig. S4), and positively correlated with serum levels of CXCL10 (Fig. 3D and Fig. S4), and CX3CL1 (Fig. 3E and Fig. S4).

**Fig. 3.**
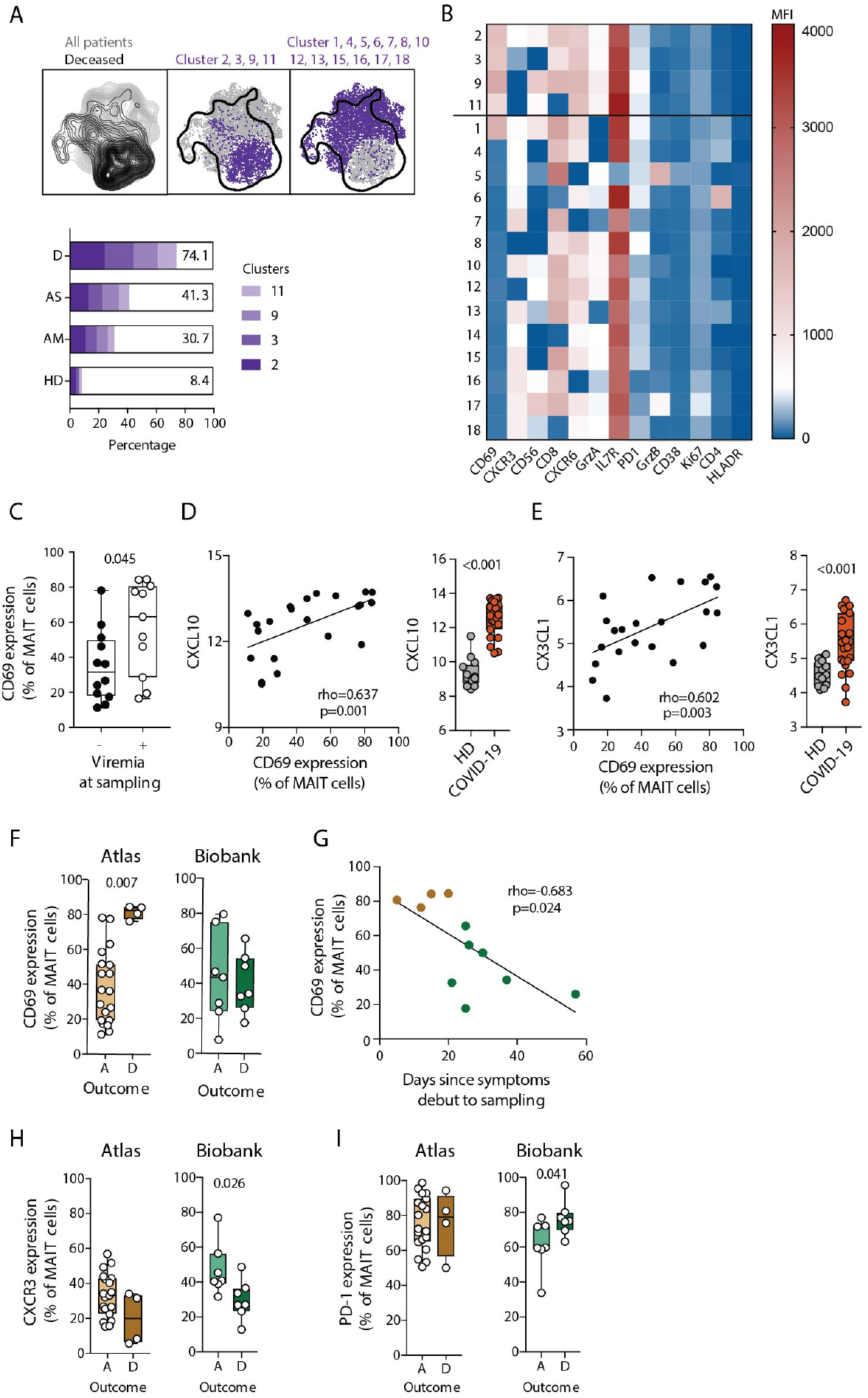
Associations between MAIT cell characteristics and COVID-19 outcome. **(A)** Top: UMAP plot of MAIT cells colored according to the outcome of the COVID-19 patients (left plot). Overlay of PhenoGraph clusters enriched in deceased (middle) or surviving (right) patients on the UMAP projection. Bottom: Bar plots summarizing the proportion of the MAIT cell clusters 2, 3, 9 and 11 for each patient group (HD, n=14; AM, n=9; AS, n=14; Deceased (D), n=4). **(B)** Heatmap of the median fluorescence intensity (MFI) of the phenotypic markers used to identify the eighteen PhenoGraph MAIT cell clusters. The black line delimitates clusters enriched (top) or not (bottom) in deceased patients. **(C)** Box plots showing CD69 expression on MAIT cells in COVID-19 patients according to their viremia status at the time of sampling (-: negative, n= 12; +: positive, n=11). **(D)** Left: Spearman correlation between CXCL10 serum level and CD69 expression on MAIT cells in COVID-19 patients (n=23). Right: box plot comparing CXCL10 serum levels in healthy donors (HD, n=14) and COVID-19 patients (n=24). **(E)** Left: Spearman correlation between CX3CL1 serum level and CD69 expression on MAIT cells in COVID-19 patients. Right: box plot comparing CX3CL1 serum levels in healthy donors and COVID-19 patients (n=23). **(F)** CD69 expression on MAIT cells in alive (A, n=19) and deceased (D, n=4) COVID-19 patients from the Atlas (left) and Biobank (right) cohorts. **(G)** Spearman correlation between CD69 expression on MAIT cells and the days since symptoms debut to sampling in deceased patients from the Atlas (brown) and the biobank (green) cohorts. **(H)** CXCR3 and **(I)** PD-1 expression on MAIT cells in alive (A) and deceased (D) COVID-19 patients from the Atlas and Biobank cohorts. **(C, D, E, F, H, I)** Each dot represents one donor. Non-parametric Mann-Whitney test was used to detect significant differences between groups. *p<0.05, **p< 0.01, ***p<0.001, ****p<0.0001. **(D, E, G)** each dot represents one donor. The spearman correlation coefficient (rho) and the associated calculated p-value (p) are indicated on the graph.

We next analyzed the markers influencing PhenoGraph clustering in relation to outcome. Strikingly, the four patients who died at hospital had significantly higher CD69 expression on their MAIT cells than patients who survived, suggesting an association between MAIT cell activation-levels and clinical outcome (Fig. 3F, left panel). To explore this pattern further, we retrospectively identified another seven deceased Intensive Care Unit (ICU) patients and seven matching ICU patients who were discharged alive (Table S2), retrieved cryopreserved Biobank PBMC samples from these patients and stained their PBMCs using the same flow cytometry panel. In this retrospective sampling of severely ill patients the association between CD69 levels and outcome was not reproduced (Fig. 3F, right panel). In comparing the first Atlas patient group with the second retrospective Biobank group, the Biobank patients were found to be sampled significantly later following symptom onset (25 vs 14 days, p < 0.001, Table S3). Plotting data from all deceased patients in both cohorts revealed an inverse correlation between MAIT cell CD69 levels and days since symptom debut to sampling (Fig. 3G), raising the possibility that high levels of MAIT cell activation early in disease may be associated with immunopathogenesis and poor outcome.

In addition to high CD69, low CXCR3 and slightly higher PD-1 expression also characterized the PhenoGraph clusters associated with poor outcome (Fig. 3B). The four patients of the Atlas cohort who died at hospital tended to have lower CXCR3 expression on their MAIT cells than patients who survived, and this pattern reached significance in the Biobank cohort (Fig. 3H). A similar pattern was observed with high PD-1 expression on MAIT cells, which was associated with poor outcome in the Biobank cohort (Fig. 3I). Together, these findings suggest that activation and chemokine receptor expression in MAIT cells is associated with disease severity and may be associated with clinical outcome of COVID-19 disease.

### Recovery of the MAIT cell compartment in COVID-19 convalescent individuals

Some chronic viral diseases are associated with partial loss of MAIT cells in circulation, which may be persistent with failure to recover when viremia is suppressed or cleared by treatment (*35-37*). To determine the ability of MAIT cells to recover after COVID-19, we analyzed peripheral blood samples drawn from patients recovering from mild disease (MC) (n=23), and from patients in the convalescent phase after severe COVID-19 (SC) (n=22), within one to six weeks from resolution of disease (Table S1). Compared with the pooled Atlas AM and AS patients groups (all acute = A), both MC and SC groups had MAIT cell frequencies, determined as percentage of T cells, similar to those of the HD group (Fig. 4A). The phenotypic flow cytometry characteristics of MAIT cells in the HD, A, MC, and SC groups were analyzed using principal component analysis (PCA) (Fig. 4B). The convalescent groups were largely overlapping with the HD group and distinct from the acute patients, where PC1 separated the all acute group based on expression of activation markers. Some convalescent individuals appeared distinct from the HD group, and were separated primarily by the relative representation of CD8^+^ and DN MAIT cell subsets contributing to PC2. Direct comparison of groups indicated that CD69 levels were largely normalized in convalescent patients (Fig. 4C). In contrast, CXCR3 levels were still suppressed, in particular in the SC group, raising the possibility that low CXCR3 expression may be a persistent alteration in MAIT cells post-COVID-19 (Fig. 4D). CD38 and other activation markers tended to normalize (Fig. S5). Together, these results indicate that MAIT cells recover in circulation within weeks of resolution of COVID-19 symptoms, although some phenotypical perturbations still persist.

**Figure 4.**
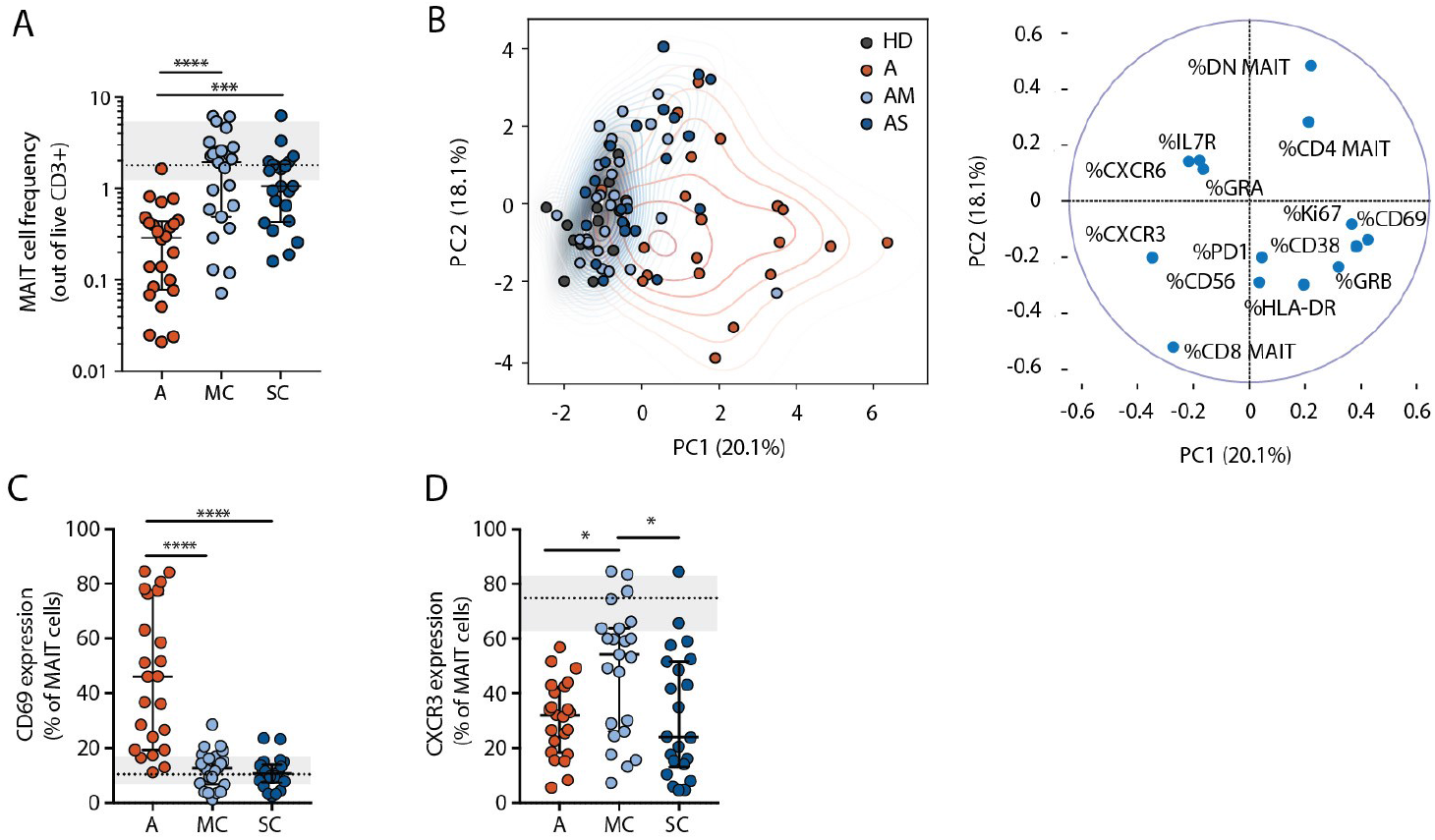
Recovery of MAIT cells in convalescent COVID-19. **(A)** Graph showing MAIT cell frequency (median ± IQR) in acute (A, n=23), moderate convalescent (MC, n=23), and severe convalescent (SC, n=22) COVID-19 patients. MAIT cell frequency (median ± IQR) in healthy donors (HD, n=14) is shown in gray. **(B)** Left: PCA plot showing the distribution and segregation of MAIT cells between healthy donors (gray dots) acute (red dots n=23), mild convalescent (light blue, n=23) and severe convalescent (dark blue, n=22) COVID-19 patients. Right: PCA biplot representing the influence of each parameter on principal component 1 (PC1) and 2 (PC2). **(C)** Graph illustrating CD69 and **(D)** CXCR3 expression on MAIT cells in acute, MC and SC patient groups. Expression in HD (median ± IQR) is shown in gray. **(A, C, D)** Each dot represents one patient. Non-parametric Kruskal-Wallis test and Dunn’s post-hoc test were used to detect significant differences between the acute and convalescent groups. *p<0.05, **p<0.01, ***p<0.001, ****p<0.0001.

## Discussion

MAIT cells play a significant role in the immune defence against microbial infections in mucosal barriers via TCR-mediated recognition of MR1-presented riboflavin metabolites. However, MAIT cells also function as innate sensors of inflammation and viral infection via activation by cytokines such as IL-18 and IFNα (12). We here found that MAIT cells are highly activated in COVID-19, and decline sharply in numbers in circulation already in moderate disease in a manner correlated with serum levels of CCL20, CXCL11, CCL28 and IL-17C. Notably, the MAIT cell lymphopenia was more profound than that observed for conventional CD8 and CD4 T cells, and other unconventional T cell subsets. Findings in two separate scRNAseq data sets indicated that MAIT cells are highly enriched among T cells infiltrating the airways during COVID-19.

MAIT cell activation levels (CD69^high^) were associated with detectable plasma viremia and correlated with increased serum levels of CXCL10 and CX3CL1. MAIT cell activation may be associated with downregulation of CXCR3, as indicated by the inverse correlation between CD69 and CXCR3. The CXCR3^low^ phenotype seems more stable and was seen also in the mucosal scRNAseq data. Notably, both these MAIT cell phenotypes (CD69^high^ and CXCR3^low^) were associated with poor clinical outcome among COVID-19 patients as assessed in the present cohort. One should bear in mind however that the number of patients studied here is small, and these MAIT cell phenotypes should not be interpreted as predictive biomarkers of poor outcome. Nevertheless, these associations and patterns support a model where MAIT cell activation may be part of the broader virus-driven type I IFN response (38), and recruitment to the airways may reflect a possible active participation of MAIT cells in the response against SARS-CoV-2 replication and later in perpetuation of the pathological inflammatory process in COVID-19. The interpretation of preferential recruitment of MAIT cells to the inflamed airways gains further support from the distinct tissue homing profile of MAIT cells with expression of several relevant chemokine receptors, such as CCR6, CXCR6, CCR5 and CXCR3 (*34*). This notion has further support in the literature, as activation and homing to sites of inflammation is consistent with observations in obesity and diabetes (*39*), multiple sclerosis (*40*), and in inflammatory bowel disease (*41*). Moreover, recent findings from bacterial pathogenesis indicate that the innate-like responsiveness of MAIT cells is important in initiation and amplification of the hyper-inflammatory response to bacterial superantigens (*42, 43*).

A striking pattern in the scRNAseq data was that the MAIT cell cluster displayed an IL-17A biased profile and was the main T cell cluster expressing *IL17A* transcript. Previous studies have shown that mucosal tissue MAIT cells have an IL-17 biased pro-inflammatory profile with less IFNγ expression (*23, 44*). Thus, interpretation of these findings in the context of the current MAIT cell immunobiology literature suggests a model where MAIT cells are activated in response to SARS-CoV-2 infection, preferentially recruited to the inflamed airway mucosa, and contribute to tissue inflammation during COVID-19 disease.

Persistent loss of MAIT cells can have detrimental long term consequences for immune defense against microbial disease and immune homeostasis at barrier sites (*45, 46*). Thus, it is interesting and promising that MAIT cells seem to recover in convalescent COVID-19 patients, even within weeks from resolution of symptoms. This pattern is compatible with recruitment to the airways rather than physical loss of MAIT cells during active disease. It is possible that, as inflammation is resolved, the MAIT cells are again released to circulation. With regard to the phenotypical MAIT cell characteristics, most seem to normalize in convalescence. However, CXCR3 expression remains suppressed and some convalescent donors have a disturbed balance of CD8^+^ and DN MAIT cell subsets. The CXCR3^low^ phenotype is more stable than the more transient CD69^high^ phenotype. Overall, the relatively rapid recovery of the MAIT cell compartment bodes well for the ability of these individuals to control future microbial infections, although this topic will require dedicated studies.

Loss of detectable CXCR3 expression in MAIT cells seems to be linked to activation as indicated by the inverse correlation with CD69 expression. However, a range of other chemokine receptors might be involved in recruitment of MAIT cells to inflamed airways. This includes CXCR6, which is consistently expressed by MAIT cells in healthy donors, and largely remains expressed in COVID-19 albeit with somewhat lower expression in patients with reduced MAIT cell frequencies (Fig. 2F). It is interesting to note that the gene encoding CXCR6 is one of six genes located within the 3p21.31 locus strongly associated with severe COVID-19 in a recent genome wide association study (*47*). The role of CXCR6, as well as of CCR5 and CXCR4, in MAIT cell recruitment in COVID-19, and in COVID-19 in general, is currently unclear and should be topics of future investigation.

The current study presents evidence for a role of MAIT cells in COVID-19 disease, which is distinct from that of adaptive conventional T cells. However, the study at the same time has several limitations. COVID-19 is heterogeneous in its presentation with a wide range of disease severity, symptoms and kinetics of disease. While the patient groups studied here were all well-defined, they still do not reflect the full complexity of the disease. Furthermore, the cross-sectional design does not capture the full disease dynamics. In particular, sampling very early following infection or symptom debut would be valuable in future studies. Nevertheless, despite these limitations the current study suggests a role for MAIT cells in COVID-19 and opens new avenues to be explored for better understanding of the immunopathogenesis of this disease.

## Materials and Methods

### Patient characteristics

SARS-CoV-2-infected patients 18 to 78 years old (n=69) with acute COVID-19 disease admitted to the Karolinska University Hospital, Stockholm, Sweden, or followed up in convalescent phase, were recruited to the study. Healthy controls (n=14) were SARS-CoV-2 IgG seronegative at time of inclusion. Patients in the Karolinska COVID-19 Immune Atlas cohort were sampled 5-24 days after symptom debut and 0-8 days after hospital admission in the acute phase. Those classified as having acute moderate (AM) COVID-19 disease (n=9) had oxygen saturation of 90-94% or were receiving 0.5-3 L/min of oxygen at the time of inclusion. Patients with acute severe (AS) COVID-19 (n=15) required either >10 L/min of oxygen at sampling or invasive mechanical ventilation, and were treated in one of the Karolinska University Hospital ICUs or a high dependency unit. For both AM and AS groups, patients with current malignant disease or ongoing immunomodulatory treatment prior to hospitalization were excluded. Samples from individuals in the convalescent phase after severe disease (SC) (n=22) were collected 42-58 days after disease onset, corresponding to 3-21 days after resolution of symptoms (100% were antibody-seropositive for SARS-CoV-2). Samples obtained from individuals in the convalescent phase after mild disease (MC) (n=23), were collected 49-64 days after disease onset, corresponding to 25-53 days after resolution of symptoms. Samples from the COVID-19 Biobank cohort were PBMC from ICU patients cryopreserved in liquid nitrogen, and thawed immediately before staining (n=14). The severity of the disease was also graded with NIH Ordinal Scale (48), and SOFA score at the sampling (49). For detailed clinical information, see Tables S1 and S2. The study was approved by the Swedish Ethical Review Authority and all patients gave informed consent.

### Flow cytometry and antibodies

The following antibodies were used for staining: CD69-BUV395 (clone FN50), CD38-BUV496 (clone HIT2), CD56-BUV737 (clone NCAM16.2), CD3-BUV805 (clone UCHT1), CD14-V500 (clone M5E2), CD19-V500 (clone HIB19), Vα24-BV750 (clone L243), CD161-PE-Cy5 (clone DX12), GrzB-AF700 (clone GB119) from BD Biosciences, PD1-BV421 (clone EH12.2H7), CD8-BV570 (clone RPA-T8), IL7R-BV605 (clone A019D5), CXCR3-BV650 (clone G025H7), CD4-BV711 (clone OKT4), HLA-DR-BV785 (clone L243), Ki-67-AF488, GrzA-PercCP-Cy5.5 (clone CB9), TCRγδ-PE/Dazzle564 (clone B1), Vα7.2-PE-Cy7 (clone 3C10), CXCR6-AF647 (clone K041E5) from Biolegend. PE conjugated 5-OP-RU loaded human MR1 tetramer (NIH tetramer core facility) was used for the identification of MAIT cells. LIVE/DEAD™ Fixable Near-IR Dead Cell Stain Kit (ThermoFischer) was used for the staining of dead cells. Cells were first stained with the hMR1-5-OP-RU tetramer for 30 min at room temperature (RT) prior to extracellular staining for another 20 min at 4°C in PBS 2 mM EDTA and 2% FBS. After washing, cells were fixed and permeabilized in BD Cytofix/Cytoperm™ Fixation/Permeabilization Kit (BD Biosciences) for 30 min at 4°C and stained intracellularly in 1X BD Perm/Wash buffer (BD Biosciences) for 30 min at 4°C. After washing, cells were fixed for two hours at RT in 1% paraformaldehyde before acquisition. Samples were acquired on a BD FACSymphony A5 flow cytometer (BD Biosciences) and analysed with FlowJo software version 10.6.2 (FlowJo, LLC). Single-stained compensation beads (BD Biosciences) were used to calculate compensation matrix prior to sample acquisition. Stainings were performed on freshly isolated (Atlas and convalescent cohorts) or cryopreserved (Biobank cohort) PBMCs.

### Serum proteomics

Sera were evaluated for soluble factors using proximity extension assay technology (Olink AB, Uppsala). All sera were heat-inactivated (56°C for 30 min) before analysis.

### Trucount

Absolute counts in whole blood were assessed by flow cytometry using BD Multitest™ 6-color TBNK reagents in association with BD Trucount™ tubes (BD Biosciences) following the manufacturer’s instructions. Samples were fixed 2 h at RT in 1% PFA before acquisition on the BD FACSymphony (BD Biosciences). CD3^+^ T cells were gated out of CD45^+^CD14^-^CD15^-^CD19^-^cells and the number of events obtained was used to determine the absolute CD3 counts as follow: (Number of CD3^+^ events acquired x number of beads per tubes) / (number of beads events acquired x sample volume in µL). The absolute count of each T cell subset analyzed was subsequently calculated using their frequencies out of total CD3^+^ cells.

### Single-cell RNAseq analysis

Publicly available scRNAseq datasets were analyzed using standard Seurat (3.2.0) workflow. Briefly, RDS files from the Chua et al. (https://doi.org/10.6084/m9.figshare.12436517) and Liao et al. (http://cells.ucsc.edu/covid19-balf/nCoV.rds) datasets were downloaded and subsetted on non-B lymphocyte clusters, then clustered and projected on UMAP. The clusters containing MAIT cells were identified based on expression of *KLRB1, DPP4, IL7R*, and *SLC4A10*, as well as *TRAV1-2* transcripts when available in the data. MAIT cell frequency among T cells was then estimated as the percentage of cells belonging to the MAIT containing cluster among all cells annotated as T cells.

### UMAP and PhenoGraph analysis

FCS3.0 files exported from BD FACSDiva software were imported into FlowJo software and automated compensation matrix was generated using the acquired single-stained compensation beads. To generate the UMAP, samples were first down-sampled using the FlowJo plugin Downsample (V2.0.0), barcoded with patient group and patient outcome, and concatenated. The FlowJo plugin UMAP (V2.2) was run on the resulting FCS file using the default settings (Distance function = Euclidean, nearest neighbors: 15 and minimum distance: 0.5) and including all the compensated parameters and FSC and SSC measurements. For cluster identification, the FlowJo plugin PhenoGraph (V2.4) was run on the resulting UMAP using the default settings (nearest neighbors K=30) and including the following parameters: CD69, CD38, HLA-DR, PD1, CXCR3, CXCR6, IL7R, Ki-67, CD56, CD4, CD8, GrzB, and GrzA.

### Principal Component Analysis

PCA were performed in Python, using scikit-learn 0.22.1. Phenotypic data obtained from flow cytometry for each cell subset was normalized using sklearn.preprocessing. StandardScaler and PCA were computed on the resulting z-scores.

### Statistical analysis

Prism V7.0 (GraphPad Software) and python were used for statistical analysis. Statistically significant differences between unpaired groups were determined using non-parametric Mann-Whitney test for two group comparison or Kruskal-Wallis test followed by Dunn’s post-hoc correction when more than two groups were compared. Two parameter correlations were evaluated using the Spearman correlation. Correlation heat maps were generated in Python using the pingouin package v0.3.6 (https://pingouin-stats.org) for computing Spearman and rank-biserial correlations as well as the associated p-values.

## Data Availability

Curated flow cytometry data is available for exploration via the Karolinska COVID-19 Immune Atlas (homepage currently under construction). Other data are available on request from the corresponding author.

## List of Supplementary Materials

**Table S1**. Demographic and clinical characteristics of the acute COVID-19 Atlas cohort, healthy donors, and convalescent patients.

**Table S2**. Demographic and clinical characteristics of the COVID-19 Biobank cohort patients.

**Table S3**. Demographic and clinical data comparison between the acute COVID-19 Atlas cohort and the Biobank cohort.

**Fig. S1**. Gating strategy, MAIT cell abundance and alteration of soluble factors in COVID-19 patients.

**Fig. S2**. Phenotypic alterations of peripheral blood MAIT cells and other T cell subsets in COVID-19 patients.

**Fig. S3**. Transcriptional profiling of the MAIT cell compartment in the upper respiratory tract.

**Fig. S4**. Associations and correlations of MAIT cell count and phenotype with clinical parameters, cytokine and chemokine levels in serum.

**Fig. S5**. Phenotypic comparison of peripheral blood MAIT cells in acute and convalescent COVID-19 patients.

## Acknowledgments

We express our gratitude to all donors, health care personnel, study coordinators, administrators, and laboratory managers involved in this work. The MR1 tetramer technology was developed jointly by Dr. James McCluskey, Dr. Jamie Rossjohn, and Dr. David Fairlie, and the material was produced by the NIH Tetramer Core Facility as permitted to be distributed by the University of Melbourne.

## Authorship Contributions

JKS, TP, JBG, KM, JE, MB, JK, and KS designed experiments. TP, JBG, JE, TS, APP, and ORB performed experiments. TP, JBG, AP, TK, and JKS analyzed data. KS, SA, EF, OR, LIE, ANT, NB, HGL, and MB provided critical material. JKS supervised the work. JKS, TP and JBG wrote the paper. All authors reviewed and approved the final manuscript.

